# Assessing the Accuracy of CGM Metrics: The Role of Missing Data and Imputation Strategies

**DOI:** 10.1101/2025.02.13.25322196

**Authors:** Simon Lebech Cichosz, Thomas Kronborg, Stine Hangaard, Peter Vestergaard, Morten Hasselstrøm Jensen

## Abstract

**Aim:** This study aims to evaluate the accuracy of continuous glucose monitoring (CGM)-derived metrics, particularly those related to glycemic variability, in the presence of missing data. It systematically examines the effects of different missing data patterns and imputation strategies on both standard glycemic metrics and complex variability metrics.

**Methods:** The analysis modeled and compared the effects of three types of missing data patterns—missing completely at random (MCAR), segmental gaps, and blockwise gaps—with proportions ranging from 5% to 50% on CGM metrics derived from 14-day profiles of individuals with type 1 and type 2 diabetes. Six imputation strategies were assessed: data removal, linear interpolation, mean imputation, piecewise cubic Hermite interpolation, temporal alignment imputation (TAI) and random forest-based imputation.

**Results:** A total of 933 14-day CGM profiles from 468 individuals with diabeteswere analyzed. Across all metrics, the coefficient of determination (R^2^) improved as the proportion of missing data decreased, regardless of the missing data pattern. The impact of missing data on the agreement between imputed and reference metrics varied depending on the missing data pattern. To achieve high accuracy (R^2^ > 0.95) in representing true metrics, at least 80% of the CGM data was required. While no imputation strategy fully compensated for high levels of missing data, simple removal and TAI outperformed others in certain scenarios.

**Conclusion:** This study highlights the significant impact of missing data and imputation strategies on CGM-derived metrics, particularly glycemic variability and time below range (TBR) estimates. The findings underscore the necessity of rigorous data handling practices to ensure reliable assessments of glycemic control and variability.

## Introduction

Continuous Glucose Monitoring (CGM) systems have significantly advanced diabetes management by offering near-continuous, real-time tracking of glucose levels [1–4]. CGM systems provide a rich source of data that allows for comprehensive assessment of glycemic control, beyond what traditional metrics like HbA1c can offer. By delivering insights into daily glucose fluctuations, CGM metrics such as time-in-range (TIR), time above range (TAR), and time below range (TBR) have become integral to evaluating the effectiveness of treatment strategies and guiding personalized care for individuals with diabetes [5]. CGM metrics provide critical information on not only average glucose levels but also the variability and risk of glycemic excursions, which are key to understanding short- and long-term outcomes. Furthermore, CGM data can be leveraged to develop clinical predictive models for diagnostics and prognostics purposes [6–11].

In clinical practice and research, CGM data is typically assessed over 14-day periods, with prior studies indicating that up to 30% missing data during this timeframe does not significantly impact the accuracy of metrics like TIR when assessing overall glycemic control [12–14]. These findings have reinforced the utility of CGM for short-term glucose monitoring and decision-making, even when data is incomplete. Despite this progress, a critical gap remains in understanding the broader implications of missing data, particularly its effect on more nuanced metrics like glycemic variability (GV). Metrics such as coefficient of variation (CV), mean amplitude of glycemic excursions (MAGE), and standard deviation of glucose are increasingly recognized for their role in predicting complications and tailoring therapeutic interventions [15], yet the impact of data incompleteness on these measures has not been thoroughly investigated.

Missing data is an inevitable challenge in CGM due to sensor malfunctions, signal loss, or patient adherence issues, and it introduces uncertainty in the calculation of these metrics [16]. The reliability of CGM data in the presence of missing data hinges on how effectively these gaps are addressed[17]. Imputation strategies—ranging from simple methods like mean imputation or linear interpolation to more sophisticated machine learning approaches—can influence the estimated values of both standard and advanced glycemic metrics. However, the relative performance of these imputation strategies in preserving the accuracy of glycemic variability estimates remains largely unexplored. This lack of evidence presents a significant limitation, as the choice of imputation strategy could potentially introduce bias or misrepresent glycemic patterns, leading to suboptimal clinical decisions.

The primary aim of this study was to assess the accuracy of CGM-derived metrics, particularly those related to glycemic variability, in the presence of missing data. We systematically evaluated the influence of different missing data patterns and imputation strategies on both standard glycemic metrics and more complex variability metrics. By doing so, we aim to determine the extent to which missing data impacts these metrics and identify the imputation strategy that best mitigate this impact. Our findings will provide insights into how CGM data should be handled in research and clinical practice, enhancing the reliability of glycemic variability metrics used in diabetes management and potentially guiding the development of standardized protocols for managing missing CGM data.

## Methods

The study methodology involved modeling and comparing the effects of three distinct types of missing data, with missing data proportions ranging from 5% to 50%, on the estimation of CGM metrics derived from 14-day profiles. Additionally, six imputation strategies for handling missing data were evaluated. A schematic representation of the methodological framework is provided in Figure 1.

**Figure 1.**
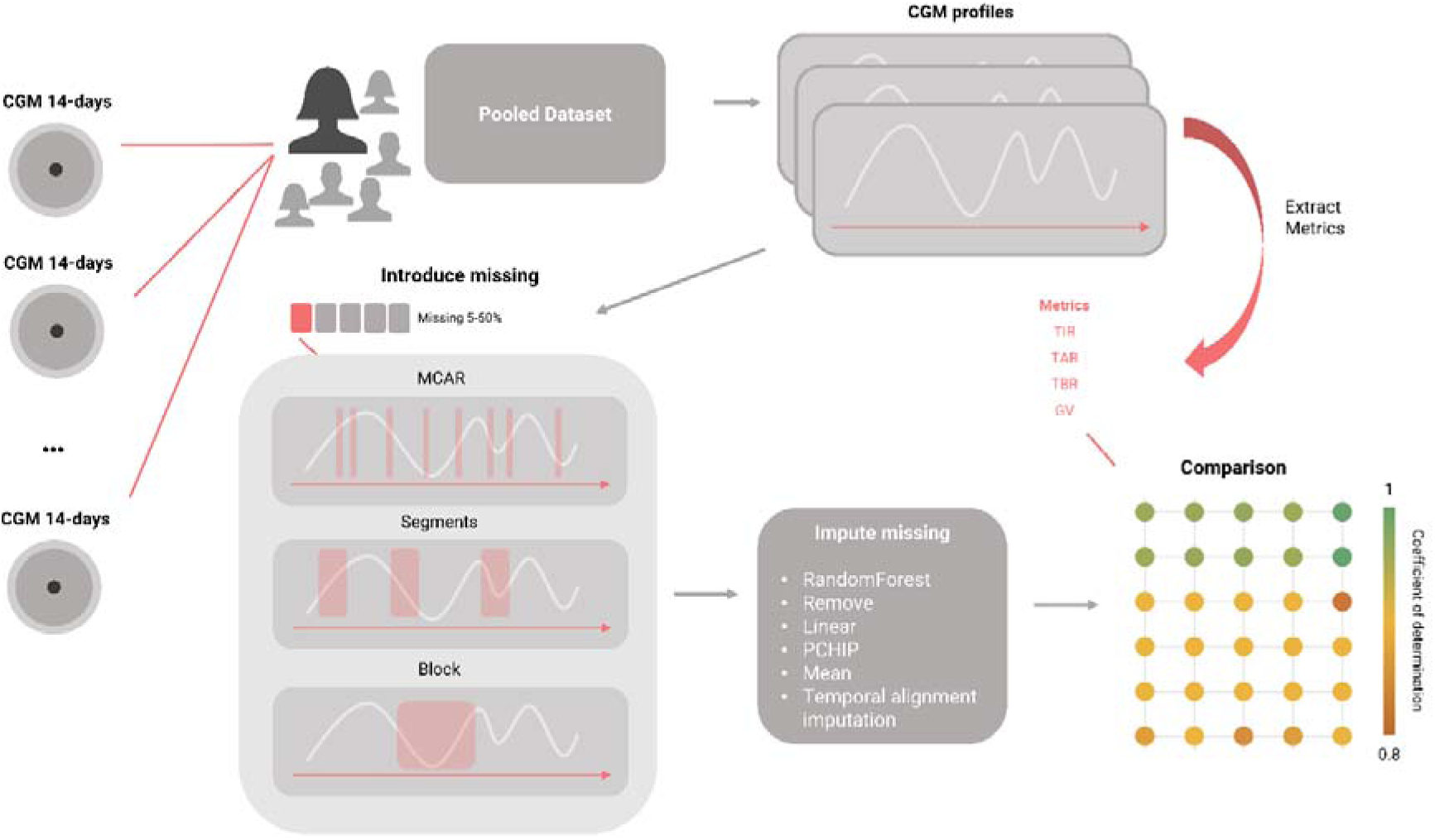
Schematic representation of the methodological framework. Continuous glucose monitoring (CGM) 14-day profiles with less than 1% missing data are pooled to form the dataset for analysis. True CGM metrics are computed from the complete profiles. Various methods are then applied to introduce missing values, followed by different imputation techniques. The metrics derived from the imputed profiles are subsequently compared to the true metrics.

### Study sample

This analysis utilized data from two studies on type 1 and type 2 diabetes;

the Diabetes TeleMonitoring of Patients in Insulin Therapy (DiaMonT) trial [18] (NCT04981808), an open-label randomized controlled trial involving 331 individuals with insulin-treated type 2 diabetes (T2DM). Conducted over three months at two sites in Denmark, participants were randomized 1:1 to either a telemonitoring intervention group or a usual care control group. The telemonitoring group employed a CGM (Dexcom G6), a connected insulin pen, an activity tracker, and smartphone applications, with hospital staff providing remote monitoring and periodic telephone contact. The usual care group used a blinded CGM for the first and last 20 days of the trial and a connected insulin pen (without smartphone applications) throughout the trial.
The Insulin-Only Bionic Pancreas Pivotal Trial (IOBP) (NCT04200313) [19]: This multicenter, randomized controlled trial evaluated the efficacy of an at-home closed-loop insulin delivery system compared to standard care. The study included participants aged 6 to 79 years with type 1 diabetes (T1D). Over a period of up to 13 weeks, participants in the intervention group used the iLet Bionic Pancreas system with the Dexcom G6 CGM system for insulin administration, while the control group continued with standard care without the closed-loop system.

### Preprocessing

For this analysis, CGM data from both the intervention and control groups were evaluated for inclusion. CGM data for each participant were divided into 14-day segments, a timeframe recommended for assessing glycemic metrics [12,13,20]. Segments were included if they contained at least 99% of the expected data points, based on a sampling interval of five minutes and the segment duration. This stringent inclusion criterion was established to ensure accurate estimation of glycemic metrics prior to modeling the effects of various missing data patterns and imputation strategies.

### CGM metrics

For calculating CGM metrics we utilized an open-source tool, Quantification of Continuous Glucose Monitoring (QoCGM), designed for comprehensive CGM data analysis within the MATLAB environment [21]. We included a range of commonly reported from basis descriptive statistics, time-in-ranges, glycemic risk indicators and glycemic variability metrics to characterize the CGM data for each 14-day segment. These metrics provide foundational insights into the central tendency, general glycemic control, dispersion, and fluctuations in glucose levels over time:

- Mean Glucose: The average glucose level over the monitoring period.
- Standard Deviation (SD): A measure of glucose variability around the mean.
- Coefficient of Variation (CV): The standard deviation normalized by the mean, expressed as a percentage.
- Time In Range (TIR) (70-180 mg/dL) [12]: Percentage of time within the target range, indicating overall glycemic control.
- Time In Tight Range (TITR) (70-140 mg/dL) [22]: Percentage of time within a narrower target range, reflecting tighter glucose control.
- Total Time Below Range (TBR) (<70 mg/dL) [23]: Percentage of time in hypoglycemia.
- Total Time Above Range (TAR) (>180 mg/dL) [12]: Percentage of time in hyperglycemia.
- Continuous Overall Net Glycemic Action (CONGA) [24]: This metric measures glucose variability over different time intervals. We calculated CONGA for 2-hour, 6-hour, and 24- hour periods (CONGA 1H, CONGA 2H, CONGA 6H, and CONGA 24H), reflecting short-term to daily fluctuations in glucose levels.
- Mean Amplitude of Glycemic Excursions (MAGE) [25]: MAGE captures the average magnitude of significant glucose swings, both increases and decreases, by focusing on excursions that exceed one standard deviation from the mean. It is a widely used indicator of glycemic variability and the likelihood of large glucose fluctuations.
- Mobility: This metric assesses the signal mobility by measuring the variance of glucose changes (differences between consecutive glucose readings) relative to the overall variance of the glucose signal. Higher mobility indicates greater variability in glucose levels. [26]
- Distance Traveled per Minute (DTpM) [27]: DTpM quantifies the total amount of glucose fluctuation over time by summing the absolute changes in glucose levels and normalizing by the total duration of monitoring. This metric provides a rate of glucose change, highlighting periods of rapid fluctuations.
- Glucose Management Indicator (GMI) [28]: The GMI provides an estimate of average glucose levels by applying a formula that correlates with HbA1C values. This metric offers a standardized measure of overall glycemic control and helps in evaluating the long-term efficacy of diabetes management.
- Overall Glycemic Risk Index (GRI) [29]: A composite metric measuring the glycemic control quality.

### Missing data modeling

To assess the impact of missing data on CGM metrics derived from 14-day profiles, we introduced three patterns of missingness to the dataset: missing completely at random (MCAR), segment-based gaps, and block-based gaps. The proportions of missing data were systematically varied from 5%, 10%, 20% 30%, 40% to 50% to simulate different scenarios of data loss in CGM datasets. Figure 2 exemplify the three different approaches to missingness in one profile.

**Figure 2.**
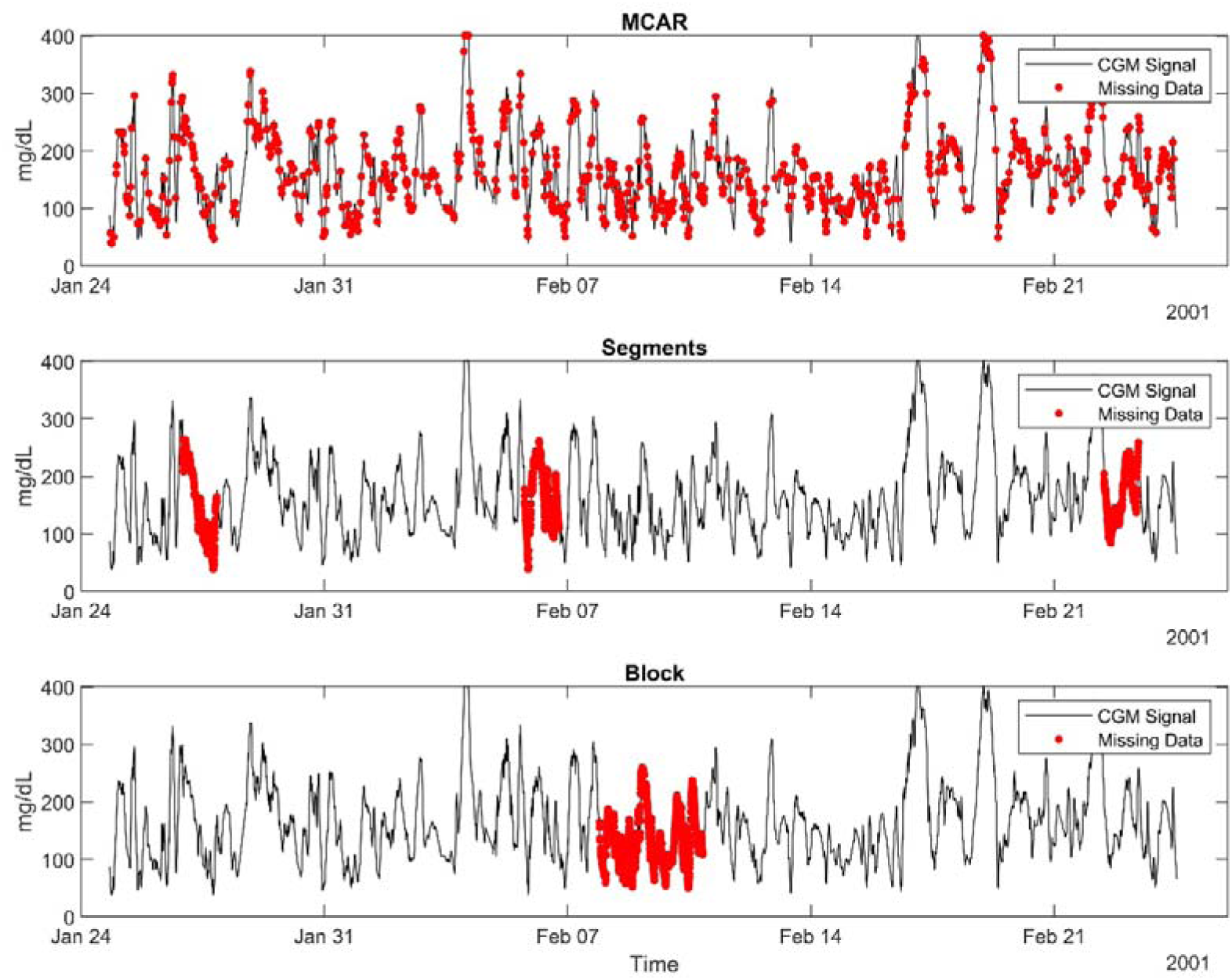
Illustration of the three methods (MCAR, Segments, Block) used to introduce missing data into the continuous glucose monitoring (CGM) profiles. In this example, 10% of the data is missing. The MCAR (Missing Completely at Random) method introduces missing values randomly across the entire dataset. The Segments method introduces missing values within three randomly selected time segments. The Block method introduces missing values within a single continuous time period. The figure showcases an synthetics CGM signal with the purpose of illustrating the methods.

*MCAR*: Data points were randomly removed throughout the time series, mimicking unpredictable loss of CGM readings.

*Segments*: Missing data were introduced in intermittent segments, reflecting scenarios such as temporary device malfunctions.

*Blocks*: Larger, contiguous blocks of data were removed, simulating extended periods of device disconnection, poor adherence or failure.

### Imputation Strategies

To handle missing data effectively, we applied and evaluated six distinct handling techniques, each representing a different approach to reconstructing the CGM profiles [30–34]. These methods are outlined below:

*Removal*: In this approach, periods containing missing data were removed entirely from the analysis. This is one of the commonly used methods to handle missing data.

*Linear Interpolatio*n: A deterministic method that estimates missing data by fitting a straight line between two known data points immediately preceding and following the gap. This method is computationally simple and effective when missing segments are short and the data exhibits relatively stable trends over time.

*Mean Imputation*: In this method, missing data are replaced with the mean of the observed values within the dataset or a specified time window. While simple and computationally efficient, mean imputation assumes uniformity in the data and may underestimate variability, potentially biasing some versatile metrics.

*Piecewise Cubic Hermite Interpolation (PCHIP)*: This spline-based technique fills gaps using cubic Hermite polynomials, preserving the shape and monotonicity of the surrounding data. PCHIP is particularly useful when capturing the non-linear trends often present in CGM data, as it prevents unrealistic oscillations between interpolated points.

*Random Forest-based Imputation (RF)*: This machine learning-based approach uses the Random Forest algorithm to estimate missing data. It captures complex, non-linear relationships, making it potentially useful for datasets with intricate patterns of variability.

*Temporal Alignment Imputation (TAI):* This proposed method addresses missing data by replacing them with the median of corresponding values recorded at the same hour on different days for each individual patient.

Each of these methods was applied across the three missingness patterns (MCAR, segments, and blocks) and evaluated over varying proportions of missing data (5%–50%).

### Comparative Analysis

The reliability of imputation strategy or simple removal was assessed by comparing the metrics derived from imputed profiles against the original (complete) dataset. The coefficient of determination (R^2^) was calculated for each metric across all scenarios to quantify the agreement and performance of imputation strategies. There is currently no established clinical consensus on the acceptable degree of agreement for such analyses. For the purposes of this study, we defined the levels of agreement as follows: "Good alignment" was classified as an R^2^ > 0.95 "Alignment" as an R^2^>0.90, and "Acceptable alignment" as an R^2^ > 0.80. The assessment was conducted on the general level of R^2^ of all metrics and on individual metrics to nuance the understanding.

## Results

A total of 933 14-day CGM profiles from 468 individuals (Type 1+2) with diabetes were included in the analysis. Hence, 453 14-day CGM profiles from 231 patients with type 2 diabetes mellitus (T2DM) undergoing insulin therapy were included in the analysis. Of the 331 patients from the DiaMonT trial, 100 were excluded due to insufficient data completeness (<99% data over the 14- day period). In addition, 480 14-day CGM profiles from 237 patients with type 1 diabetes mellitus were included in the analysis. Of the 440 patients from the IOBP study, 203 were excluded due to insufficient data completeness (<99% data over the 14-day period).

For all metrics, the coefficient of determination (R^2^) improved as the proportion of missing data decreased, irrespective of the missing data pattern. The effect of missing data proportions on the agreement between imputed and reference measures varied according to the pattern of missing data, as shown in Figure 3. For type 1 diabetes, segments and block missing patterns above 10% adversely effected the average estimate more than for type 2 diabetes profiles.

**Figure 3.**
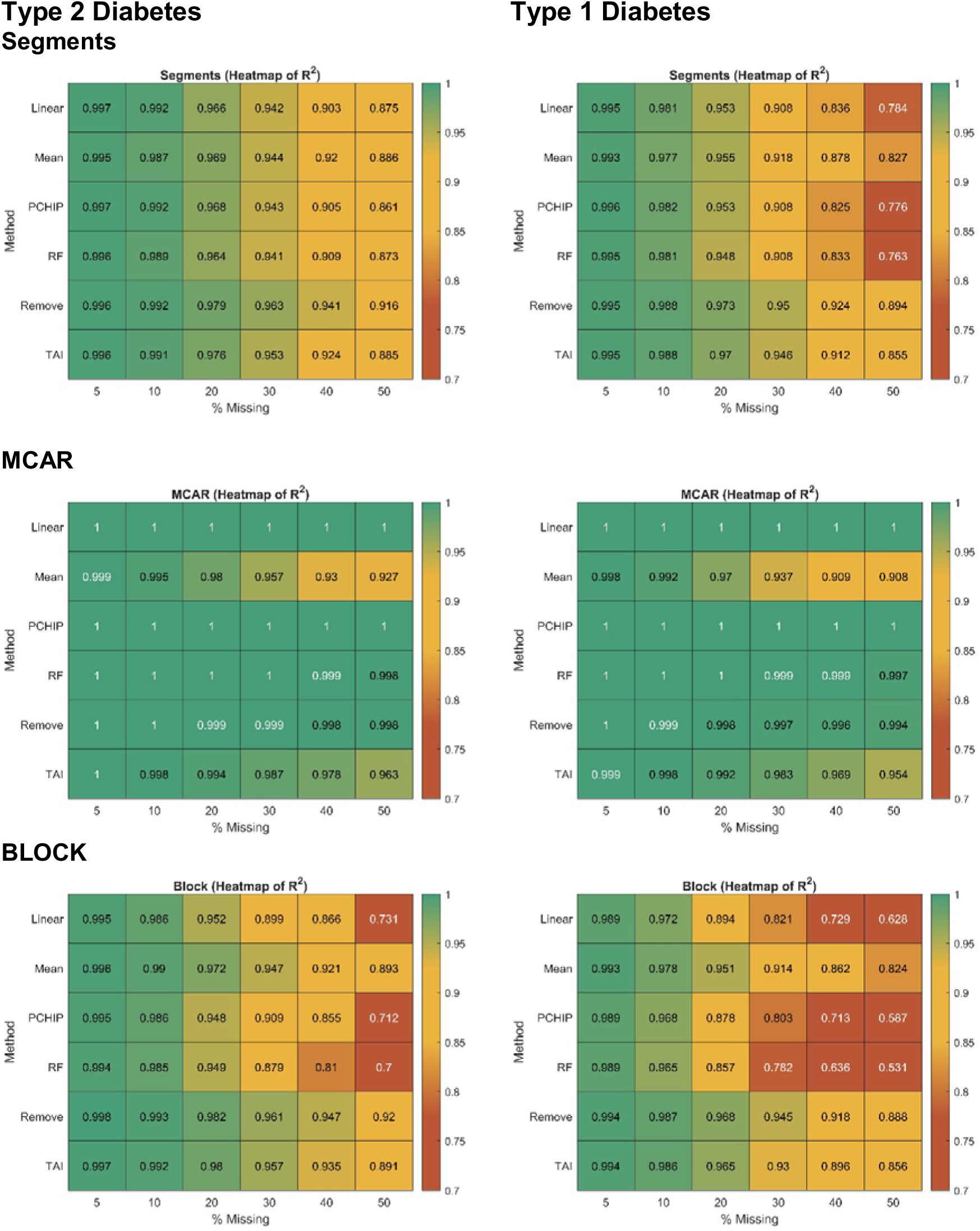
Heatmap depicting the average R^2^ values between true estimates and imputed estimates across profiles with artificially induced missing data proportions ranging from 5% to 50%, for both type 1 and type 2 diabetes.

For data missing completely at random (MCAR), R^2^ showed strong agreement across most levels of missingness and imputation strategies. Among the methods, mean imputation resulted in lower alignment compared to others, whereas simple removal, RF, PCHIP, and linear interpolation achieved the best alignment. For segmented and block missing patterns, a low proportion of missing data (5–20%) yielded good alignment across removal, TAI, mean. However, at higher proportions of missing data (20–50%), removal and TAI outperformed other methods in alignment under these patterns. The average R^2^ across all metrics, however, does not fully capture the effects of missingness and imputation strategies on individual metrics. Figure 4 + 5 illustrates the alignment of individual metrics under 10%, 30%, and 50% missing data, with additional detailed data provided in Supplementary Data 1–6.

**Figure 4.**
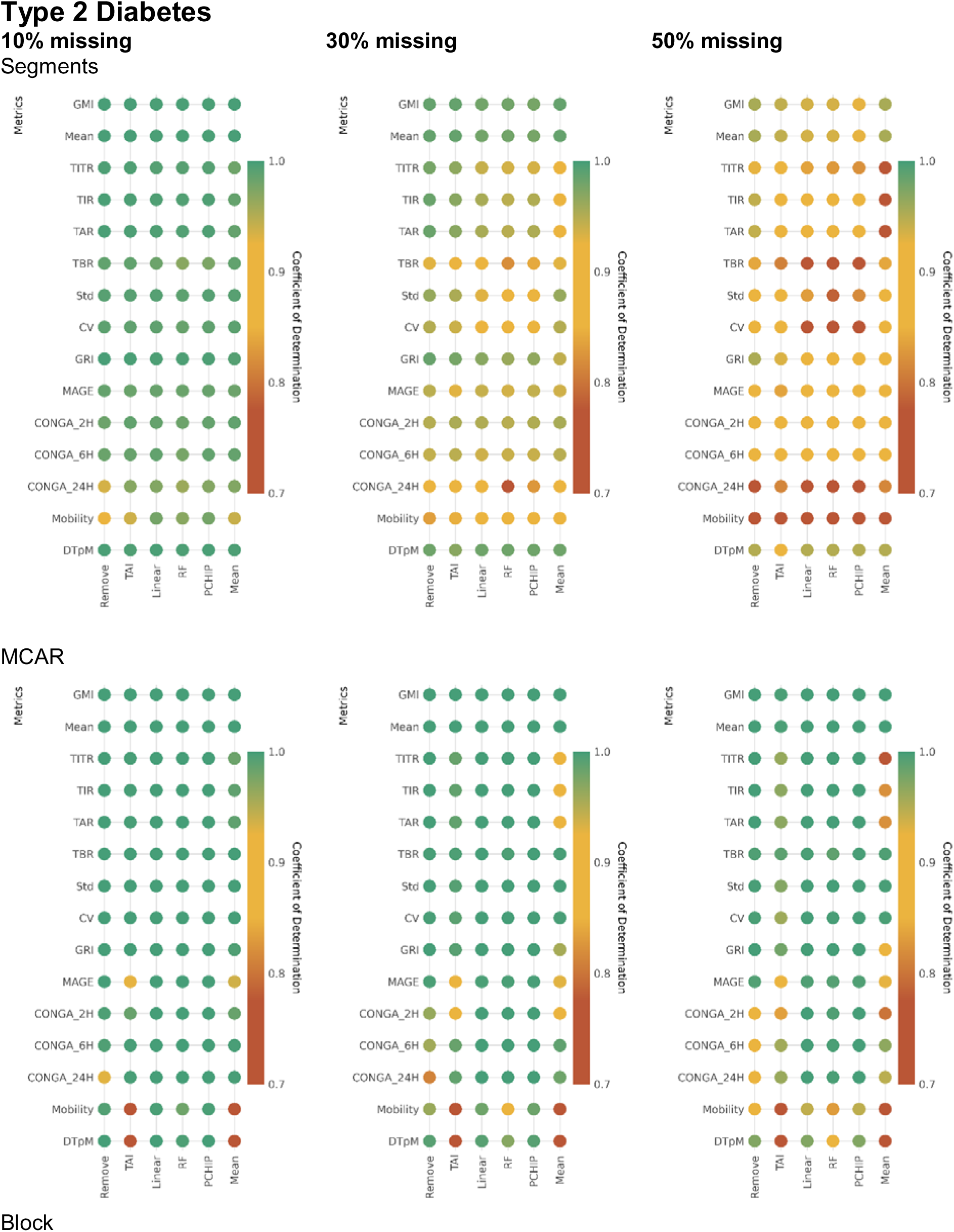
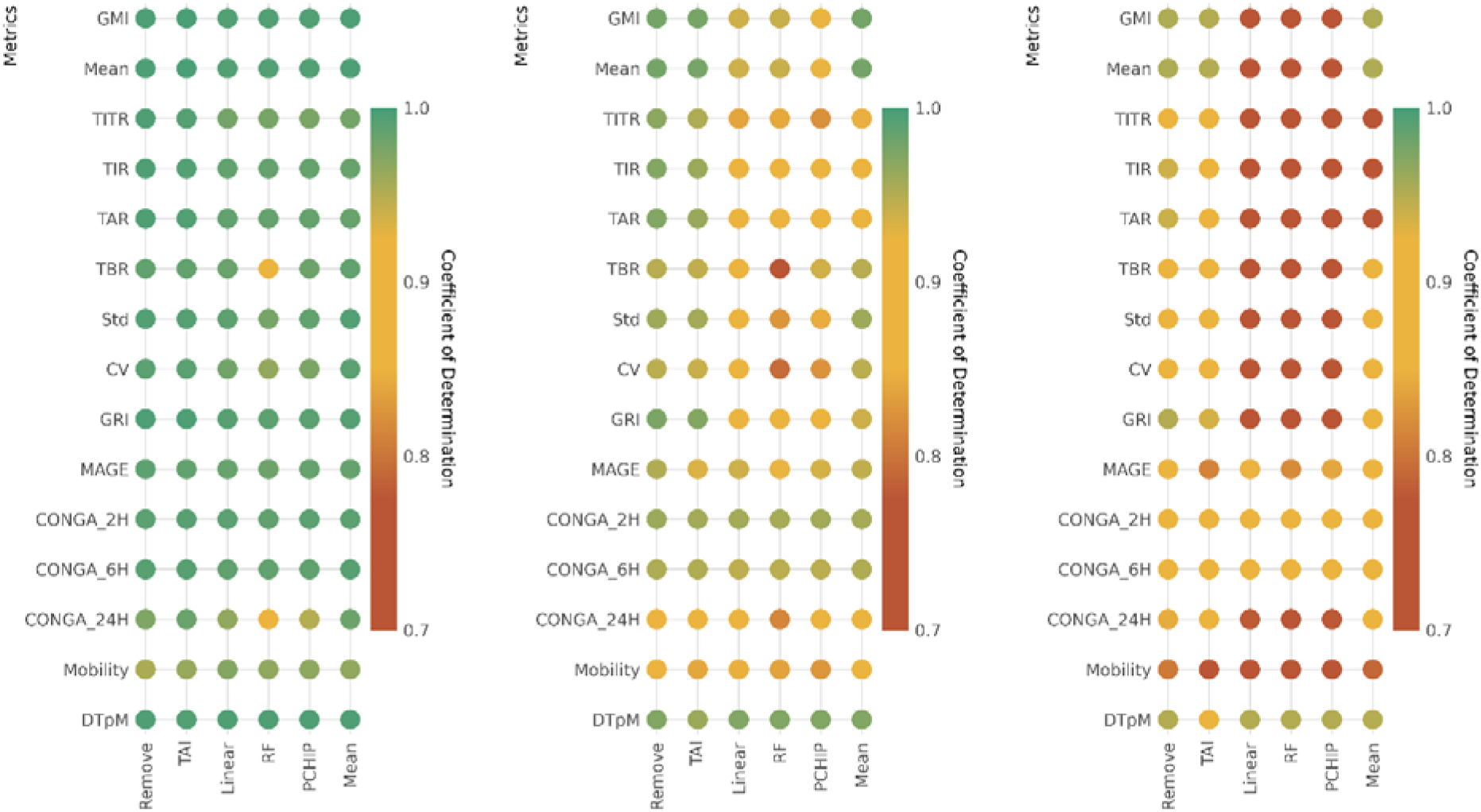
Heatmap depicting the R^2^ values between true estimates and imputed estimates for each CGM metric across profiles with artificially induced missing data proportions ranging from 5% to 50%, for type 2 diabetes.

**Figure 5.**
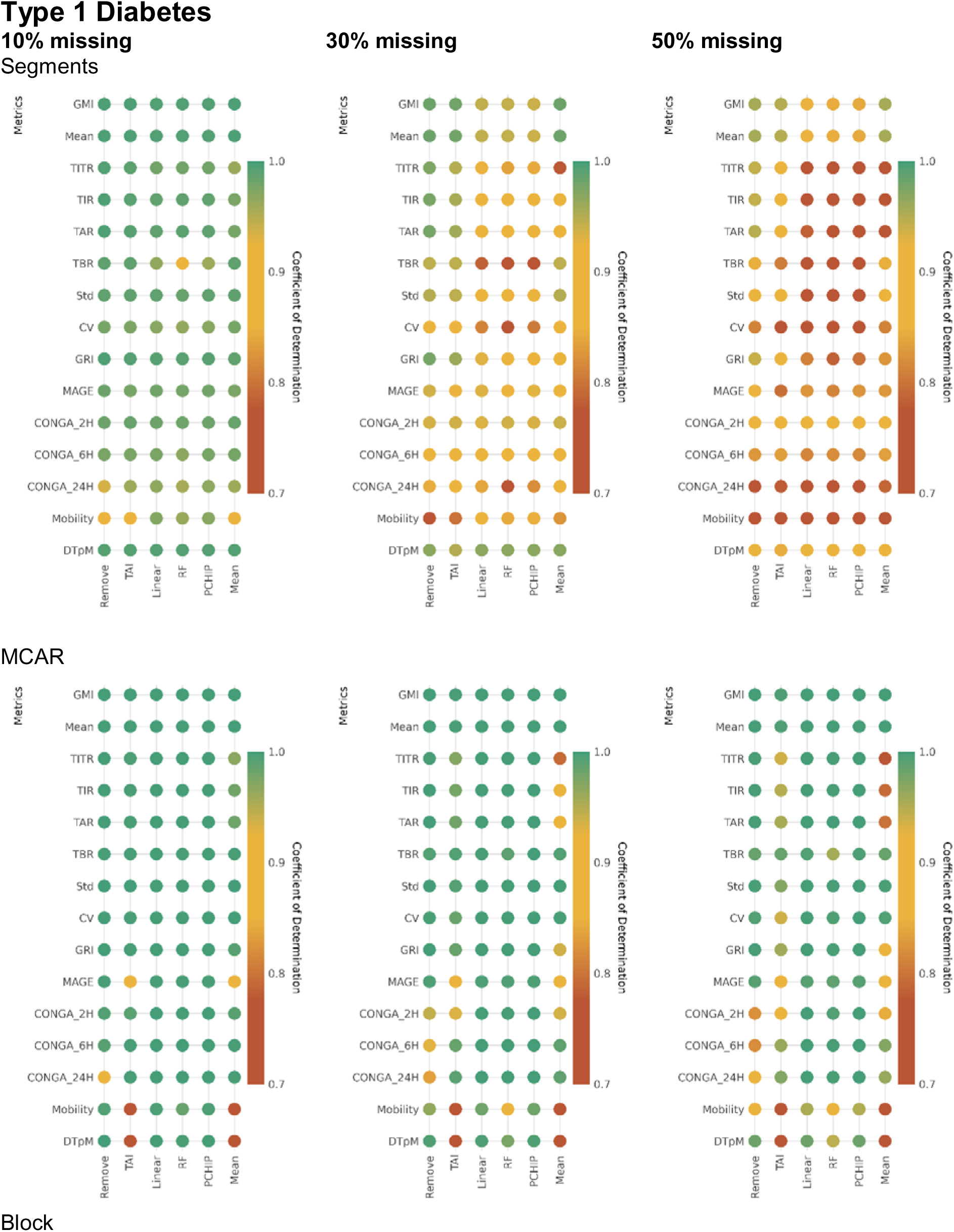
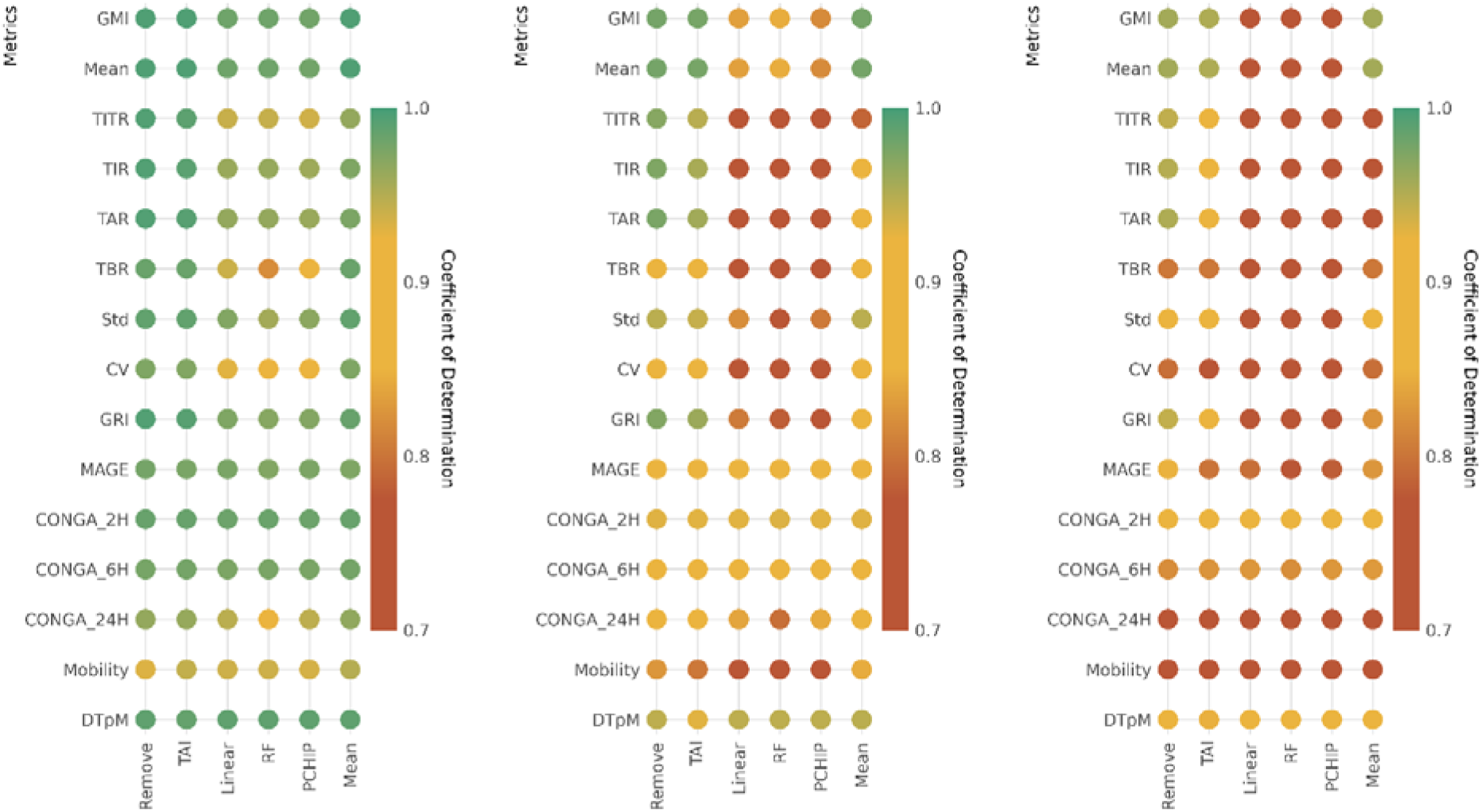
Heatmap depicting the R^2^ values between true estimates and imputed estimates for each CGM metric across profiles with artificially induced missing data proportions ranging from 5% to 50%, for type 1 diabetes.

For MCAR patterns, high alignment between imputed CGM data and reference estimates was observed, except for certain metrics when using mean imputation or TAI. Metrics such as DTpM, mobility, MAGE, and TITR were particularly negatively affected by mean imputation.

Under segmented missing patterns, good alignment was observed for missing data ≤20%. At higher levels of missing data, alignment was generally preserved for most metrics, with removal and TAI demonstrating a slight advantage.

For block missing patterns, strong alignment was observed for missing data ≤20%. At higher levels of missing data (<50%), alignment was achieved for most metrics, although alignment deteriorated significantly at 50% missing data for most metrics. Notably, removal and TAI improved the estimates for several metrics under high missingness (30-50%).

## Discussion

This study systematically evaluated the impact of missing CGM data on standard glycemic metrics and glycemic variability metrics, highlighting how different imputation strategies influence the reliability of these metrics under various missing data patterns. The findings provide important insights for both clinical and research applications of CGM, where missing data is a common challenge.

The results demonstrate that the proportion and pattern of missing data significantly affect the reliability of CGM metrics. Metrics generally exhibited strong agreement with reference values when missingness was ≤20%, regardless of the pattern. However, as missingness increased (30–50%), the alignment between imputed and reference metrics varied depending on both the pattern of missing data, metric and the imputation strategy.

The study highlights those low levels of missingness (≤20%) are generally acceptable, regardless of imputation strategy. However, as missingness increases, careful consideration is required to select appropriate imputation strategy.

Advanced Metrics (e.g., MAGE, GV, mobility) are more sensitive to the choice of imputation method, with removal, TAI and mean imputation underperforming in MCAR cases and preforms better in more realistic cases with missing patterns related to segments or block patterns.

However, statistical patterns of missing data in CGM have not been thoroughly characterized in the literature. One study points towards that missingness is missing not at random (MNAR), with an overrepresentation of missing data during the night [35].

For High Missingness (>30%) - For segmented and block patterns, removal and TAI, while simplistic, demonstrated reasonable performance for some metrics, suggesting it may serve as a practical fallback. However, its use must be used with caution to avoid introducing biases in GV and time dependent metrics. Also worth mentioning, recommended clinical practice is a lower (<30%) missingness in 14-day CGM profiles [12].

In comparison with previous published work, Smith et al [16] recently investigated the effects of missing data patterns on commonly used metrics (time in ranges, mean and CV) in children and adolescents with type 1 diabetes. Their findings showed that across all missing data patterns, R^2^ values decreased as the proportion of missing data increased. However, as the size of contiguous blocks of missing data grew, the impact of missing data on the agreement between measures became more pronounced. For a 14-day CGM dataset to reliably represent percentage TIR, at least 70% of the data should be available, covering a minimum of 10 days. Measures with skewed distributions, such as percentage TBR and CV, were more sensitive to missing data compared to less skewed measures. Also, Akturk et al. [36] showed that under MCAR patterns, a substantial amount of data loss can be tolerated without significantly compromising the accuracy of CGM-derived glycemic metrics. Braber et al. [37] reported the effect of missingness on clinical interpretation from patients with type 1 and 2 diabetes using the FreeStyle Libre sensor. They concluded that 30% missingness from a 14-day profile is generally acceptable, however for reliable interpretation of TBR in profiles with a low GMI, data loss should be< 10%. We extended the understanding of the impact of missing data by incorporating various interpolation strategies and evaluating a comprehensive range of CGM metrics across both type 1 and type 2 diabetes populations. Kuang et al. [38] recently examined imputation strategies for four-week CGM profiles collected from non-diabetic pregnant women. The study aimed to assess the bias introduced across various CGM summary metrics when using complete case analysis, where missing data are excluded, as well as several single imputation, multiple imputation, and machine learning-based approaches. The findings revealed that complete case analysis and hot-deck imputation exhibited the best performance based on the evaluated CGM metrics.

Our results underscore the need for standardized protocols to manage missing CGM data. The variability in metric reliability under different missingness scenarios suggests that one-size-fits-all imputation strategies are insufficient. Instead, tailoring the approach based on missingness patterns and metric priorities is important.

The primary limitations of this study include the assessment of a limited set of missingness scenarios and imputation strategies. Additionally, the scenarios evaluated were based on data from individuals participating in a clinical trial, which may affect the generalizability of the findings. Caution should also be exercised when extrapolating these results to CGM profiles with durations beyond 14 days and CGM metrics not included in this analysis. Furthermore, the analysis focused exclusively on profiles obtained from the Dexcom G6 sensor, limiting the applicability of the findings to other sensor systems without further validation.

Future work should expand these findings by exploring the performance of advanced machine learning-based imputation methods, which may offer improved accuracy for complex metrics under high missingness. Additionally, further validation in diverse populations and with different CGM systems would enhance generalizability.

Overall, this study provides evidence on the impact of missing data and imputation strategies on CGM-derived metrics, particularly glycemic variability metrics and TBR estimates. The findings emphasize the importance of careful handling of CGM data to ensure accurate assessments of glycemic control and variability, ultimately supporting more informed clinical decision-making and research insights.

## Data Availability

The data utilized (DiaMont study) in this study are not publicly available due to the inclusion of sensitive patient information, which is subject to strict confidentiality and privacy regulations. Access to the data is restricted to ensure compliance with ethical guidelines and to protect patient privacy. Requests for additional information or collaboration may be considered on a case-by-case basis, subject to appropriate ethical approval and data-sharing agreements.

## Disclaimer

This study analyzed data from the Insulin Only Bionic Pancreas Pivotal Trial (NCT04200313), but the analyses, content and conclusions presented herein are solely the responsibility of the authors and have not been reviewed or approved by the Bionic Pancreas Research Group or Beta Bionics.

## Acknowledgments

n/a.

## Author Contributions

SLC had access to all data analyzed in this study. SLC takes responsibility for the integrity and accuracy of the study data analysis and results (conceptualization, methodology, writing - Original Draft). Other authors were involved in data collection, methodology, writing - review & editing. All authors have seen and approved the manuscript

## Funding received

none to report.

## Abbreviations

CV: Coefficient of Variation
CSV: comma-separated value
CGM: Continuous Glucose Monitoring
CONGA: Continuous Overall Net Glycemic Action
DiaMonT: Diabetes teleMonitoring of patients in insulin Therapy
DTpM: Distance Traveled per Minute
FGxP: Fasting Glucose Proxy
GMI: Glucose Management Indicator
GRADE: Glycemic Risk Assessment in Diabetes
GRI: Glycemic Risk Index
IQR: Interquartile Range
LBGI/HBGI: Low/High Blood Glucose Index
MAGE: Mean Amplitude of Glycemic Excursions
PCHIP: Piecewise Cubic Hermite Interpolating Polynomial
QoCGM: Quantification of Continuous Glucose Monitoring
SD: Standard Deviation
TAR: Time-above-range
TBR: Time-below-range
TIR: Time-in-Range
(TAI): temporal alignment imputation

## References

1 Rodbard D. Continuous Glucose Monitoring: A Review of Successes, Challenges, and Opportunities. https://home.liebertpub.com/dia. 2016;18:S23–213. doi: 10.1089/DIA.2015.0417

2 Martens T, Beck RW, Bailey R, et al. Effect of Continuous Glucose Monitoring on Glycemic Control in Patients With Type 2 Diabetes Treated With Basal Insulin: A Randomized Clinical Trial. JAMA. 2021;325:2262–72. doi: 10.1001/JAMA.2021.7444

3 Carlson AL, Mullen DM, Bergenstal RM. Clinical use of continuous glucose monitoring in adults with type 2 diabetes. Diabetes Technol Ther. 2017;19:S4–11. doi: 10.1089/DIA.2017.0024/ASSET/IMAGES/LARGE/FIGURE1.JPEG

4 Group TJDRFCGMS. Continuous Glucose Monitoring and Intensive Treatment of Type 1 Diabetes. New England Journal of Medicine. 2008;359:1464–76. doi: 10.1056/NEJMOA0805017/SUPPL_FILE/NEJM_JUVDIABRSRCHFDN_1464SA1.PDF

5 Beck RW, Bergenstal RM, Cheng P, et al. The Relationships Between Time in Range, Hyperglycemia Metrics, and HbA1c. J Diabetes Sci Technol. 2019;13:614–26. doi: 10.1177/1932296818822496/ASSET/IMAGES/LARGE/10.1177_1932296818822496-FIG4.JPEG

6 Cichosz SL, Bender C. Development of Machine Learning Models for the Identification of Elevated Ketone Bodies During Hyperglycemia in Patients with Type 1 Diabetes. https://home.liebertpub.com/dia. Published Online First: 8 March 2024. doi: 10.1089/DIA.2023.0531

7 Cichosz SL, Hejlesen O. Classification of Gastroparesis from Glycemic Variability in Type 1 Diabetes: A Proof-of-Concept Study. J Diabetes Sci Technol. 2022;16:1190–5. doi: 10.1177/19322968211015206/ASSET/IMAGES/LARGE/10.1177_19322968211015206-FIG2.JPEG

8 Fleischer J, Hansen TK, Cichosz SL. Hypoglycemia event prediction from CGM using ensemble learning. Frontiers in Clinical Diabetes and Healthcare. 2022;3:71. doi: 10.3389/FCDHC.2022.1066744

9 Cichosz SL, Jensen MH, Hejlesen O. Short-term prediction of future continuous glucose monitoring readings in type 1 diabetes: Development and validation of a neural network regression model. Int J Med Inform. 2021;151:104472. doi: 10.1016/J.IJMEDINF.2021.104472

10 Cichosz SL, Jensen MH, Olesen SS. Development and Validation of a Machine Learning Model to Predict Weekly Risk of Hypoglycemia in Patients with Type 1 Diabetes Based on Continuous Glucose Monitoring. https://home.liebertpub.com/dia. Published Online First: 12 January 2024. doi: 10.1089/DIA.2023.0532

11 Cichosz SL, Kronborg T, Jensen MH, et al. Penalty weighted glucose prediction models could lead to better clinically usage. Comput Biol Med. 2021;138. doi: 10.1016/j.compbiomed.2021.104865

12 Danne T, Nimri R, Battelino T, et al. International Consensus on Use of Continuous Glucose Monitoring. Diabetes Care. 2017;40:1631–40. doi: 10.2337/DC17-1600

13 Battelino T, Danne T, Bergenstal RM, et al. Clinical Targets for Continuous Glucose Monitoring Data Interpretation: Recommendations From the International Consensus on Time in Range. Diabetes Care. 2019;42:1593–603. doi: 10.2337/DCI19-0028

14 Cichosz SL, Jensen MH, Hejlesen O. Optimal Data Collection Period for Continuous Glucose Monitoring to Assess Long-Term Glycemic Control: Revisited. J Diabetes Sci Technol. 2023;17:690–5. doi: 10.1177/19322968211069177/ASSET/IMAGES/LARGE/10.1177_19322968211069177-FIG2.JPEG

15 Hirsch IB. Glycemic Variability and Diabetes Complications: Does It Matter? Of Course It Does! Diabetes Care. 2015;38:1610–4. doi: 10.2337/DC14-2898

16 Smith GJ, Abraham MB, De Bock M, et al. Impact of Missing Data on the Accuracy of Glucose Metrics from Continuous Glucose Monitoring Assessed Over a 2-Week Period. Diabetes Technol Ther. 2023;25:356–62. doi: 10.1089/DIA.2022.0101/SUPPL_FILE/SUPP_TABLES1.DOCX

17 Braem CIR, Yavuz US, Hermens HJ, et al. Missing Data Statistics Provide Causal Insights into Data Loss in Diabetes Health Monitoring by Wearable Sensors. Sensors 2024, Vol 24, *Page* 1526. 2024;24:1526. doi: 10.3390/S24051526

18 Hangaard S, Kronborg T, Hejlesen O, et al. The Diabetes teleMonitoring of patients in insulin Therapy (DiaMonT) trial: study protocol for a randomized controlled trial. Trials. 2022;23:1–9. doi: 10.1186/S13063-022-06921-6/FIGURES/1

19 Russell SJ, Beck RW, Damiano ER, et al. Multicenter, Randomized Trial of a Bionic Pancreas in Type 1 Diabetes. New England Journal of Medicine. 2022;387:1161–72. doi: 10.1056/NEJMOA2205225/SUPPL_FILE/NEJMOA2205225_DATA-SHARING.PDF

20 Battelino T, Alexander CM, Amiel SA, et al. Continuous glucose monitoring and metrics for clinical trials: an international consensus statement. Lancet Diabetes Endocrinol. 2023;11:42–57. doi: 10.1016/S2213-8587(22)00319-9

21 Lebech Cichosz S, Hangaard ; Stine, Kronborg T, et al. From Data to Insights: A Tool for Comprehensive Quantification of Continuous Glucose Monitoring (QoCGM). medRxiv. 2025;2025.01.01.25319870. doi: 10.1101/2025.01.01.25319870

22 Beck RW, Raghinaru D, Calhoun P, et al. A Comparison of Continuous Glucose Monitoring- Measured Time-in-Range 70–180 mg/dL Versus Time-in-Tight-Range 70–140 mg/dL. Diabetes Technol Ther. 2024;26:151–5. doi: 10.1089/DIA.2023.0380/SUPPL_FILE/SUPPL_FIGURES3.DOCX

23 Hill NR, Hindmarsh PC, Stevens RJ, et al. A method for assessing quality of control from glucose profiles. Diabetic Medicine. 2007;24:753–8. doi: 10.1111/J.1464-5491.2007.02119.X

24 Mcdonnell CM, Donath SM, Vidmar SI, et al. A novel approach to continuous glucose analysis utilizing glycemic variation. Diabetes Technol Ther. 2005;7:253–63. doi: 10.1089/DIA.2005.7.253

25 Service FJ, Molnar GD, Rosevear JW, et al. Mean Amplitude of Glycemic Excursions, a Measure of Diabetic Instability. Diabetes. 1970;19:644–55. doi: 10.2337/DIAB.19.9.644

26 Lebech Cichosz S, Kronborg ; Thomas, Laugesen E, et al. From Stability to Variability: Classification of Healthy Individuals, Prediabetes, and Type 2 Diabetes using Glycemic Variability Indices from Continuous Glucose Monitoring Data. https://home.liebertpub.com/dia. Published Online First: 8 August 2024. doi: 10.1089/DIA.2024.0226

27 Peyser TA, Balo AK, Buckingham BA, et al. Glycemic Variability Percentage: A Novel Method for Assessing Glycemic Variability from Continuous Glucose Monitor Data. Diabetes Technol Ther. 2018;20:6. doi: 10.1089/DIA.2017.0187

28 Bergenstal RM, Beck RW, Close KL, et al. Glucose management indicator (GMI): A new term for estimating A1C from continuous glucose monitoring. Diabetes Care. 2018;41:2275– 80. doi: 10.2337/DC18-1581/-/DC1

29 Klonoff DC, Wang J, Rodbard D, et al. A Glycemia Risk Index (GRI) of Hypoglycemia and Hyperglycemia for Continuous Glucose Monitoring Validated by Clinician Ratings. J Diabetes Sci Technol. 2023;17:1226–42. doi: 10.1177/19322968221085273

30 Zulj S, Carvalho P, Ribeiro R, et al. Handling Missing Data in CGM Records. IFMBE Proc. 2020;74:420–7. doi: 10.1007/978-3-030-30636-6_57/TABLES/3

31 Rehman NU, Contreras I, Beneyto A, et al. The Impact of Missing Continuous Blood Glucose Samples on Machine Learning Models for Predicting Postprandial Hypoglycemia: An Experimental Analysis. Mathematics. 2024;12:1567. doi: 10.3390/MATH12101567/S1

32 Moscardó V, Giménez M, Oliver N, et al. Updated Software for Automated Assessment of Glucose Variability and Quality of Glycemic Control in Diabetes. Diabetes Technol Ther. 2020;22:701. doi: 10.1089/DIA.2019.0416

33 Zhang XD, Zhang Z, Wang D. CGManalyzer: an R package for analyzing continuous glucose monitoring studies. Bioinformatics. 2018;34:1609–11. doi: 10.1093/BIOINFORMATICS/BTX826

34 Di J, Demanuele C, Kettermann A, et al. Considerations to address missing data when deriving clinical trial endpoints from digital health technologies. Contemp Clin Trials. 2022;113:106661. doi: 10.1016/J.CCT.2021.106661

35 Braem CIR, Yavuz US, Hermens HJ, et al. Missing Data Statistics Provide Causal Insights into Data Loss in Diabetes Health Monitoring by Wearable Sensors. Sensors 2024, Vol 24, *Page* 1526. 2024;24:1526. doi: 10.3390/S24051526

36 Akturk HK, Herrero P, Oliver N, et al. Impact of Different Types of Data Loss on Optimal Continuous Glucose Monitoring Sampling Duration. Diabetes Technol Ther. 2022;24:749– 53. doi: 10.1089/DIA.2022.0093

37 den Braber N, Braem CIR, Vollenbroek-Hutten MMR, et al. Consequences of Data Loss on Clinical Decision-Making in Continuous Glucose Monitoring: Retrospective Cohort Study. Interact J Med Res. 2024;13:e50849. doi: 10.2196/50849

38 Kuang A, Yu Y, Siddique J, et al. Imputation of Missing Continuous Glucose Monitor Data. 101177/19322968241308217. Published Online First: 31 December 2024. doi: 10.1177/19322968241308217

